# Droughts, cyclones, and intimate partner violence: A disastrous mix for Indian women

**DOI:** 10.1101/2020.06.29.20142893

**Authors:** Ayushi Rai, Anupam Joya Sharma, Malavika A. Subramanyam

## Abstract

India has reported a high prevalence of Intimate Partner Violence (IPV) against women over the years. Previous Western research have found an increased IPV risk among women in the aftermath of natural disasters, underscoring the need for such studies in India. We could not locate any study focusing on the impact of slow-onset versus rapid-onset disasters, which might have differing impacts on the vulnerable, especially on the incidence of IPV in India. Using data on ever-married women from the National Family Health Survey-4, we investigated the association of residing in districts exposed to a drought (N=31,045), and separately, to two cyclones (N=8469), with three forms of self-reported IPV against women (emotional, physical, and sexual). Survey-adjusted logistic regression models showed that exposure to cyclone was positively associated with emotional IPV (AOR: 1.59, CI: 1.20, 2.10) after adjusting for sociodemographic covariates. Although not statistically significant, exposure to cyclone was also positively associated with physical and sexual IPV, and drought with physical IPV. However, we did not find an association of drought with emotional and sexual violence. Notably, we corroborated previous findings that women from wealthier households, educated, and whose husbands had no history of alcohol consumption, were less likely to experience any form of IPV independent of the influence of other factors. These results highlight the potential increased risk of IPV following natural disasters. In a patriarchal society such as India vulnerable to climate-change, these sobering results highlight the need to prepare for the social disasters that might accompany natural disasters.

## 1. Introduction

Gender-Based Violence (GBV) is a gross human rights violation and a major public health concern impacting survivors’ physical and mental well-being. GBV is associated with serious health problems such as injuries, gynecological trauma, mental health issues including depression and post-traumatic stress disorder, adverse pregnancy outcomes, and sexually transmitted diseases including HIV/AIDS (1).

Although GBV has different forms varying with settings (2), often the violence is directed towards a woman and committed by someone close, usually an inmate partner (1). Globally, one in three ever-partnered women has experienced physical and/or sexual violence by an intimate partner (3). At the core of GBV lies unequal power relations between men and women (1) emanating from and reinforced by a deeply embedded patriarchal value system. In India, a historically patriarchal society, the prevalence of Intimate Partner Violence (IPV) was about 31% in the year 2015 (4).

The scourge of IPV usually escalates during natural disasters, which have the potential to compound pre-existing vulnerabilities by increasing the feelings of helplessness and loss of control (5). Several studies have highlighted the relationship between exposure to natural disasters (or other extreme events) and increased rates of gender-based violence or violence against women (5–13). There could be several shocks and stressors in the aftermath of natural disasters in the form of loss of life, injury, disease, damage to property, destruction of assets, loss of services, social and economic disruptions; all of which negatively affect people’s physical, mental, and social well-being. They are known to disrupt the everyday lives of the affected community by their impact on access to social networks, transportation, employment opportunities, and household resources; thereby shaking the household environment (14). Natural disasters have been shown to lead to a lack of employment opportunities, diminished social support, increased vulnerability to sex trafficking and sexual abuse, unequal access to assistance, discrimination in aid provision, and violence (13,15,16). Such stress and fear built up during the disaster warning phase could escalate further during a disaster (17). Previous study has shown that such stress (18) in the aftermath of disasters caused a feeling of inadequacy amongst men which could lead them to consume alcohol, gambling, and aggression as a way to cope with pressure. For instance, a post-Cyclone *Nargis* assessment in Myanmar found that the respondents reported an increase in alcohol consumption and 30% greater domestic violence after the disaster (19). Phillips et al (20) theorised that the reason for the increased domestic and sexual violence after disasters include threats to the male ‘provider and protector’ role, loss of control, and loss of options in sources of support for women.

In societies with limited mobility of women and social support such as India, women are more likely to experience violence (1). These societies rigidly define gender roles and have a culture tolerable to physical punishment of women and children, thus enabling violence as a way to settle interpersonal disputes. Furthermore, the strongly rooted gender norms and roles, sexist beliefs, and practices, also act as obstacles in the implementation of gender-sensitive policies (21). India’s status as one of the most disaster-prone areas in the world combined with its existing gender norms, gendered nature of labour at home and work, and gendered patterns of social structure and demography (22), increases the vulnerability of the Indian women to intimate partner violence in the aftermath of natural disasters. Despite this, we found only six studies exploring the relationship between natural disasters and intimate partner violence in India. Moreover, most of these studies were limited to understanding women’s difficulties during and after natural disasters; the post-disaster trafficking of women and girls; dowry deaths in response to weather variability; and, the gendered nature of natural disasters with a focus on post-tsunami Tamil Nadu (11,12,23–25). A recent study from India (26) found the higher prevalence of IPV after the 2004 Indian Tsunami in the four affected states. The merits of this study are the attempt to fill this gap by examining the changes in prevalence of IPV across the four Tsunami (2004) affected southern states of India in the aftermath of the disaster. However, it does not contrast the impact of slow-onset versus rapid-onset disasters on the incidence of IPV. With varied severity of natural disasters (in terms of destruction, economic loss, deaths, migration etc), their impact on the vulnerable might differ. For instance, a slow-onset disaster is likely precipitated gradually, depending on the confluence of different events and thus has a different impact than rapid or sudden-onset events which cause immediate destruction. Furthermore, Rao (26) assigns exposure to the disaster at the state-level, thus missing an opportunity to examine whether within-state variation in the exposure might have a differential impact on IPV.

With the global importance of climate change and frequent occurrence of natural disasters in India, it is crucial to analyse the influence of exposure to natural disasters on IPV. This may help design evidence-based gender-sensitive disaster recovery programs for helping the affected population. Not examining the role of natural disasters on IPV risks inaction on gender inequalities during a sensitive time and further marginalization of women. Therefore, we analyse the relationship of living in districts (sub-state divisions) exposed to two natural disasters, a drought (slow-onset) and a cyclone (rapid-onset) with violence against women by their intimate partners. Further, we examine this association under the assumption that each natural disaster might lead to different forms of IPV, namely emotional, physical, and sexual violence.

## 2. Methods

### 2.1. Data sources and the analytical sample

We used the 4^th^ round of National Family Health Survey (2015-16) for data on the sociodemographic and intimate partner violence indicators. This nationally representative survey was administered to 699,686 ever-married women aged 15-49 years. Details about the sampling, recruitment, and implementation of the survey can be found in the NFHS-4 report (4). We merged in drought-related data from the Ministry of Agriculture and Farmer Welfare, Government of India which revealed that 254 districts across 10 states were affected by drought during 2015-16. We included only these 10 states for the drought related analysis assuming that all the districts from these 10 states had equal probability of being affected by drought. After deleting observations missing data on the outcome, the analytical sample size for drought analysis was 31,045. For the cyclone-related analysis, we merged in data from The International Disaster Database (EM-DAT) widely cited in studies exploring the relationship between natural disasters and human lives (27). As per EM-DAT, 86 districts across 4 states were affected by the cyclones Phailin (2013) and Hudhud (2014). We further followed up on several government reports, Non-Governmental Organization (NGO) websites, and news articles to cross-check this list. Similar to our drought analysis, we limited our cyclone analysis to only these 4 states that were exposed to the cyclones in 2013 and 2014. After deleting observations which were missing outcome data, 8469 observations were retained for the cyclone analysis.

### 2.2. Response variables

We measured Intimate Partner Violence (IPV) along three dimensions: physical, emotional (psychological), and sexual violence (28). Responses to twelve questions from the NFHS-4 dataset (individual recode) were used to calculate the domestic violence score – 3 questions on emotional violence (whether the respondent was insulted/humiliated/threatened by the husband or partner), 7 questions on physical violence (whether the respondent was pushed/ slapped/punched/kicked/strangled/pulled hair/threatened with knife by the husband or partner), and 2 questions on sexual violence (whether the respondent was ever forced into unwanted sex/physically forced to perform sexual acts by the husband/partner). For each question about facing IPV, the responses were recorded as “never,” “sometimes,” “often,” and “yes, but not in the last 12 months.” While we coded “never” as “0,” “sometimes” and “often” were coded as “1.” Since we were focused on experiences of domestic violence after the natural disasters, we coded “yes, but not in the last 12 months” as 0. We coded emotional violence as 1 if responses to any of the questions (for emotional violence) was 1. Physical and sexual violence were dichotomized in the same manner.

### 2.3. Independent variables

#### Exposure to drought and cyclone

We created a binary variable in the drought-related dataset which assigned values of 1 and 0 to districts exposed and not exposed to the 2015-16 drought, respectively. Similarly, we created a binary variable in the cyclone-related dataset which assigned 1 and 0 respectively to districts exposed and not exposed to either of the 2013 and 2014 cyclones.

#### Covariates

We selected a range of covariates in line with previous research (26,29). We controlled for the age of the respondent (in years), level of education of the respondent, and the husband/partner (*no education*, primary, secondary, higher), sex of the household head (*male*, female), type of place of residence (rural/*urban*), quintiles of a score on household asset possession (*poorest*, poorer, middle, richer, richest), caste (*scheduled-caste*, scheduled-tribe, other-backward-classes, others), religion (*Hindu*, Muslim, Christian, Sikh, others), and husband’s alcohol consumption (yes, *no*). We also controlled for the states of residence (reference category: *Andhra Pradesh* for both cyclone and drought analyses) to account for their political and geographical diversity. The reference categories for each of the categorical variables have been italicized.

### 2.4. Data analysis

First, we calculated the prevalence of each form of IPV in the selected states by drought and cyclone history. We also computed the prevalence across levels age groups, caste, religion, and educational achievement of the respondents.

We next fitted survey-design adjusted logistic regression models to estimate the association of exposure to cyclone, and separately drought, with the three forms of IPV. For each type of IPV (emotional, physical, and sexual violence), we first fit a model (model 1) with only the primary predictor (cyclone/drought). In the second model (model 2), we additionally adjusted for all the sociodemographic covariates listed above. In addition to controlling for the covariates, our analysis accounted for the clustering due to sampling design and the domestic violence weight (d005) (4). We set *alpha* at 0.05.

All survey-adjusted logistic regression models were run in Stata version 12 (30).

### 2.5. Ethical considerations

All analyses in this study were based on publicly available data (National Family Health Survey-4). The NFHS-4 field activities were overseen by the International Institute of Population Sciences, Mumbai. A team of trained, and supervised, field assistants collected data after explaining the potential risks and receiving informed consent from the voluntary respondents. All respondents were promised anonymity and confidentiality of the data (31,32).

## 3. Results

### 3.1. Descriptive results

The mean age of the respondents in the cyclone and drought analytic datasets was 32. 64 years (SD=8.42) and 32.61 years (SD=8.16), respectively. A majority of our respondents in both analyses were older than 25 years (Table1 and Table 2). Moreover, a larger proportion of our samples belonged to the Other Backward Castes group and poorer households. A majority (∼70%) of respondents in both samples resided in rural areas. Further, the prevalence of the different forms of IPV in the groups exposed to both cyclone and drought ranged from 6.10% to 23.72% for cyclone and 4.82% to 24.15% for drought. The prevalence of different forms of IPV across diverse sociodemographic characteristics in both the analytic datasets are given in Tables 1 and 2.

**Table 1:**
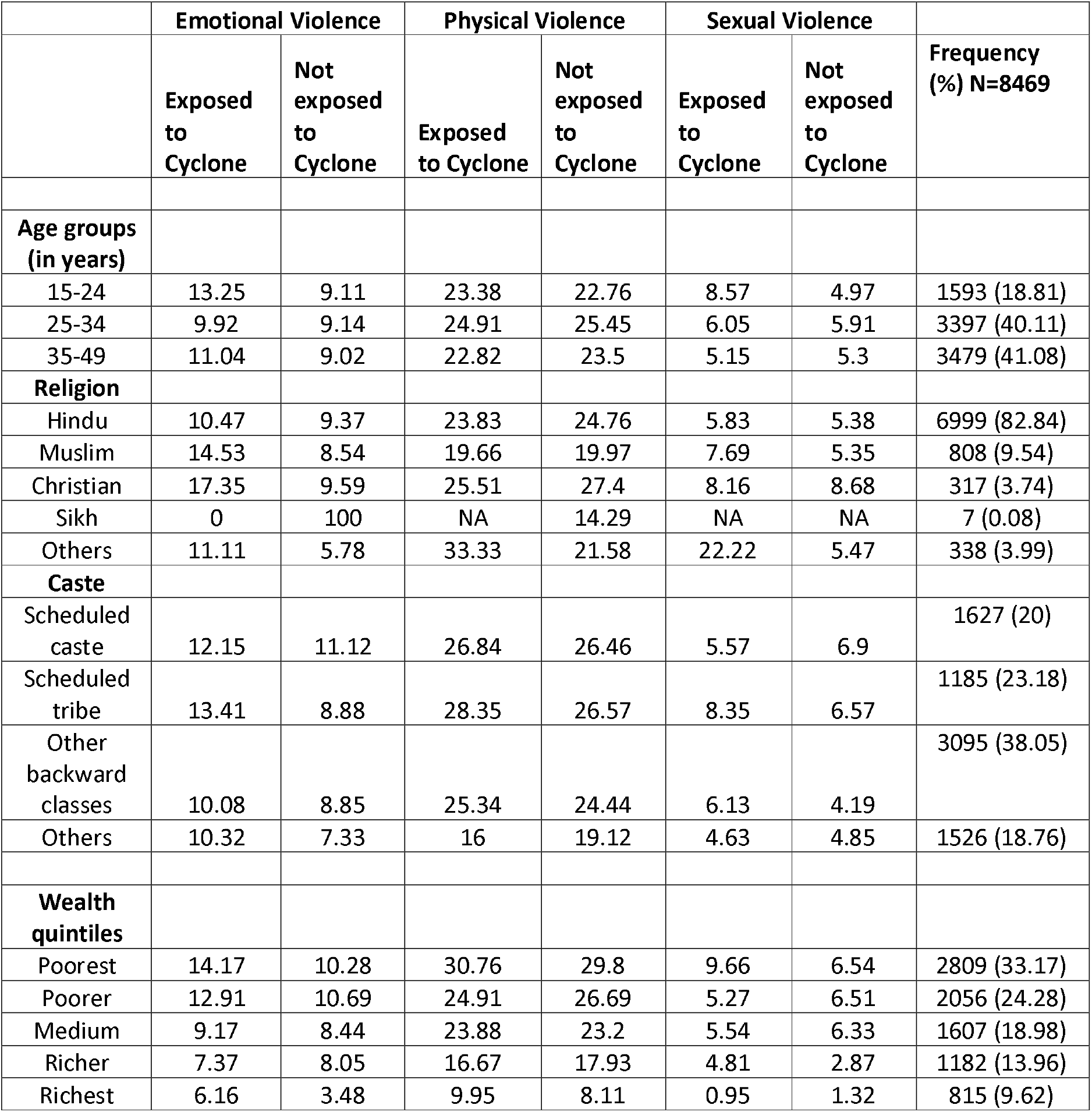
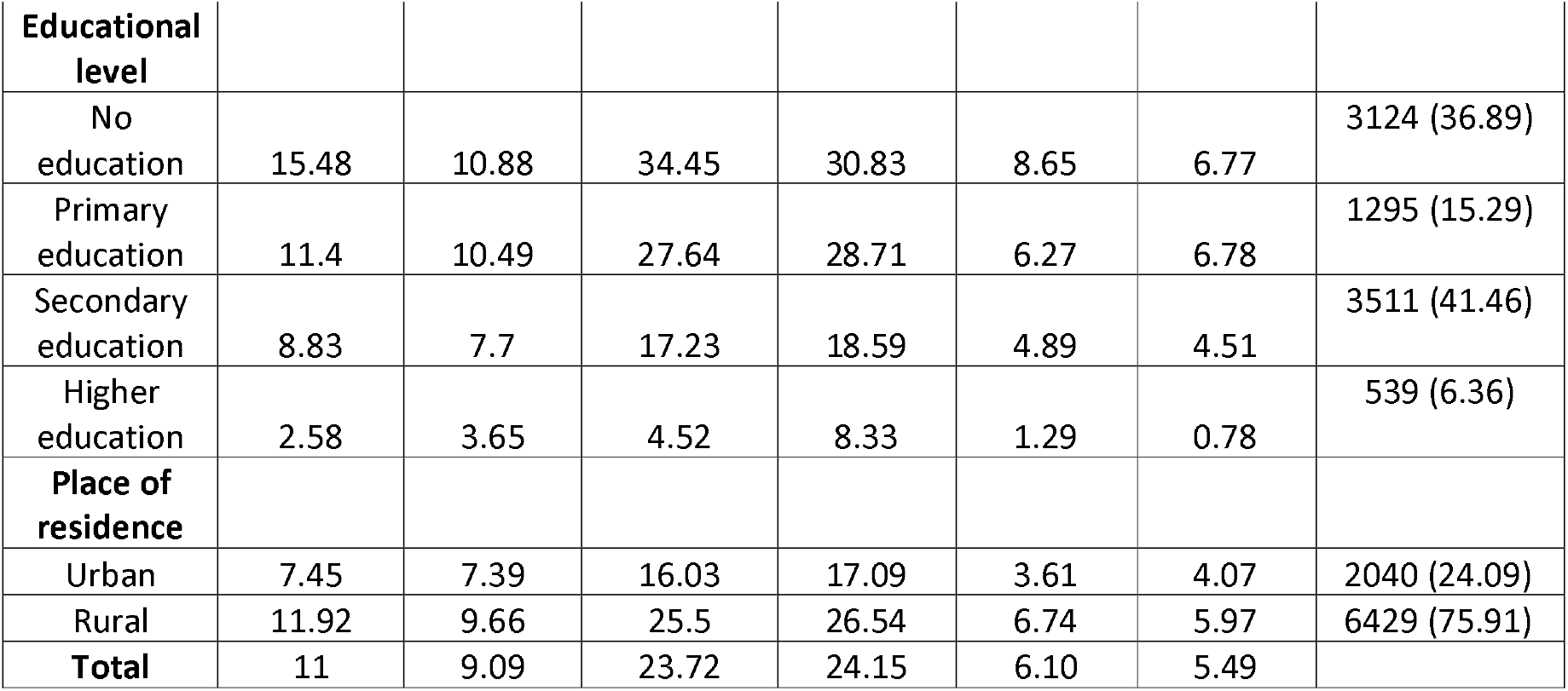
The distribution of the three forms of IPV (%) in cyclone exposed (and unexposed) groups across different sociodemographic characteristics (N=8469).

**Table 2:** The distribution of the three forms of IPV (%) in drought exposed (and unexposed) groups across different sociodemographic characteristics (N=31,045).

|  | **Emotional** |  | **Physical** |  | **Sexual** |  | **Frequency (%)**<br>**N=31,045** |
| --- | --- | --- | --- | --- | --- | --- | --- |
|  | **Exposed to Drought** | **Not exposed to Drought** | **Exposed to Drought** | **Not exposed to Drought** | **Exposed to Drought** | **Not exposed to Drought** |  |
| **Age groups (in years)** |  |  |  |  |  |  |  |
| 15-24 | 9.11 | 8.57 | 21.91 | 22.12 | 4.88 | 5.45 | 5501 (17.72) |
| 25-34 | 10.41 | 9.6 | 24.94 | 24.38 | 5.15 | 5.34 | 13,028 (41.96) |
| 35-49 | 10.54 | 9.83 | 24.32 | 21.67 | 4.45 | 4.82 | 12,516 (40.32) |
| **Religion** |  |  |  |  |  |  |  |
| Hindu | 10.3 | 9.32 | 24.48 | 23.06 | 4.84 | 5.13 | 26,774 (86.24) |
| Muslim | 10.56 | 10.34 | 21.82 | 22.07 | 4.25 | 5.7 | 3112 (10.02) |
| Christian | 10.22 | 17.65 | 24.46 | 29.41 | 7.26 | 5.88 | 440 (1.42) |
| Sikh | 4.76 | 8.51 | 21.43 | 21.28 | 4.76 | 0 | 89 (0.29) |
| Others | 6.4 | 6.92 | 19.2 | 20.77 | 4.2 | 2.31 | 630 (2.03) |
| **Caste** |  |  |  |  |  |  |  |
| Scheduled caste | 12.15 | 11.09 | 28.47 | 27.93 | 6.35 | 6.09 | 5904 (19.32) |
| Scheduled tribe | 11.19 | 11.79 | 28.5 | 30.96 | 5.56 | 7.74 | 5077<br>(16.61) |
| Other backward classes | 9.99 | 9.09 | 23.34 | 21.18 | 4.61 | 4.48 | 14,262<br>(46.70) |
| Others | 7.74 | 7.68 | 17.01 | 18.04 | 2.83 | 4 | 5309<br>(17.37) |
| **Wealth quintiles** |  |  |  |  |  |  |  |
| Poorest | 12.93 | 13.21 | 31.84 | 32.72 | 6.98 | 8.88 | 8063<br>(25.97) |
| Poorer | 11.24 | 11.13 | 26.65 | 26.86 | 4.96 | 5.6 | 7016<br>(22.60) |
| Medium | 9.88 | 10.1 | 22.63 | 24.28 | 4.64 | 5.25 | 5988<br>(19.29) |
| Richer | 8.02 | 7.82 | 18.79 | 19.35 | 3.58 | 3.78 | 5276<br>(16.99) |
| Richest | 6.04 | 5.23 | 12.57 | 11.07 | 1.86 | 2.24 | 4702<br>(15.15) |
| **Educational level** |  |  |  |  |  |  |  |
| No education | 12.66 | 11.7 | 30.5 | 28.31 | 5.98 | 6.27 | 12,487<br>(40.22) |
| Primary education | 10.74 | 10.31 | 26.21 | 27.89 | 5.53 | 6.1 | 4747<br>(15.29) |
| Secondary education | 8.44 | 7.87 | 19.37 | 17.55 | 3.95 | 4.08 | 11,338<br>(36.52) |
| Higher education | 5.26 | 4.35 | 10.24 | 10.06 | 1.6 | 2.4 | 2473<br>(7.97) |
| **Place of residence** |  |  |  |  |  |  |  |
| Urban | 9.2 | 8.53 | 19.5 | 18.14 | 3.77 | 4.05 | 8741<br>(28.16) |
| Rural | 10.59 | 10.03 | 25.8 | 25.37 | 5.19 | 5.71 | 22,304<br>(71.84) |
| **Total** | 10.23 | 9.52 | 24.15 | 22.90 | 4.82 | 5.15 |  |

### 3.2. Exposure to cyclone and IPV

Our survey adjusted single and multivariable logistic regression analyses revealed a positive association of cyclone exposure with all three forms of IPV (see Table 3). Women who were residing, versus not residing, in districts affected by the 2013/2014 cyclones had 59% higher odds (AOR: 1.59, CI:1.20, 2.10) of facing emotional violence by their husbands/partners after accounting for all the covariates. Similarly, women exposed to either of the cyclones had higher odds of facing physical (AOR: 1.24, CI: 0.97, 1.60) and sexual IPV (AOR: 1.26, CI: 0.79, 2.02) compared to women who were not exposed. Although not statistically significant, the estimates of the associations were consistent with our hypotheses. Further, our multivariable models show that women who belonged to wealthier households (rich and richest), had statistically significant lower odds of reporting emotional, physical, and sexual IPV compared to women from the poorest group, independent of all other covariates.

**Table 3:**
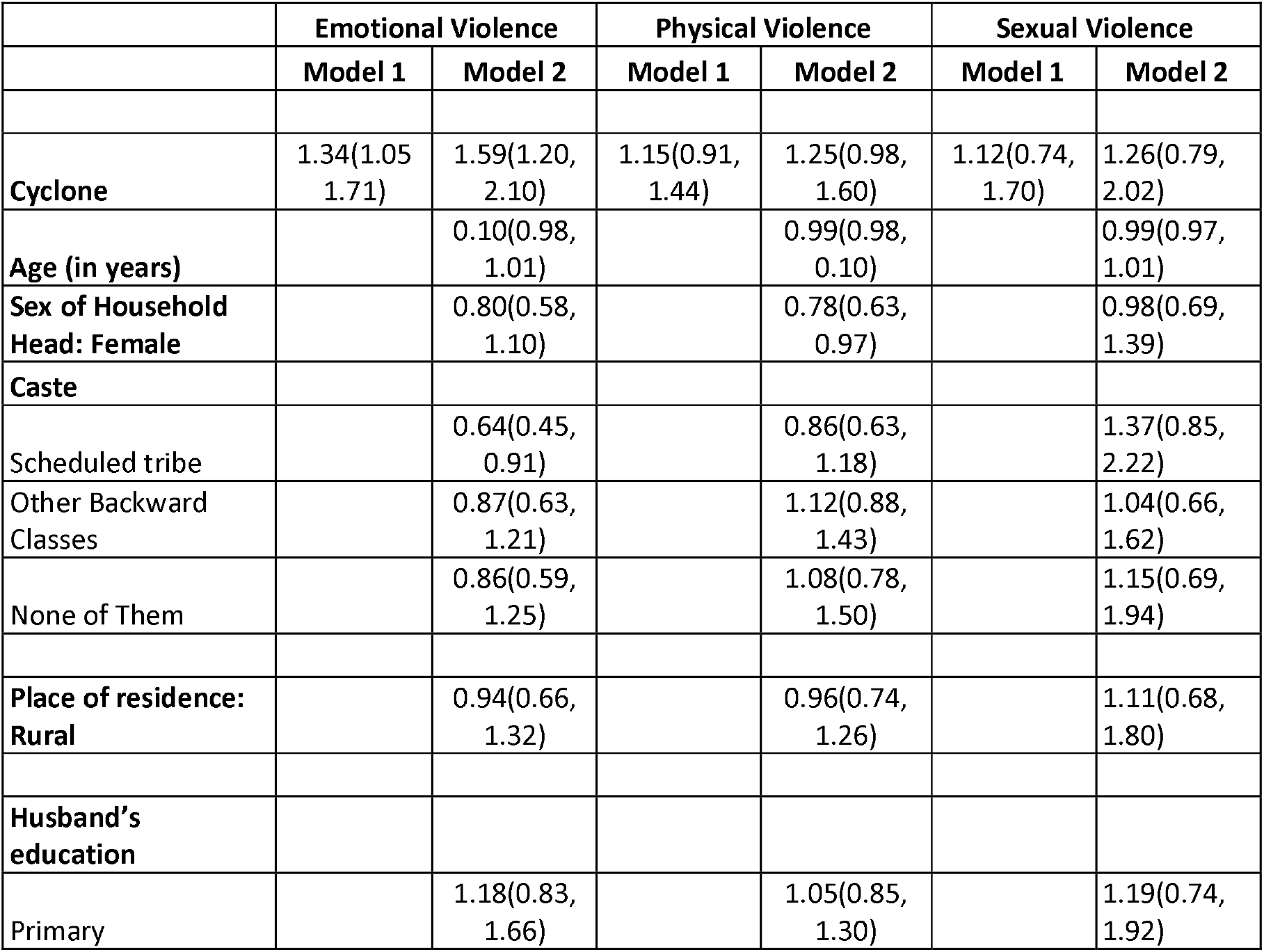
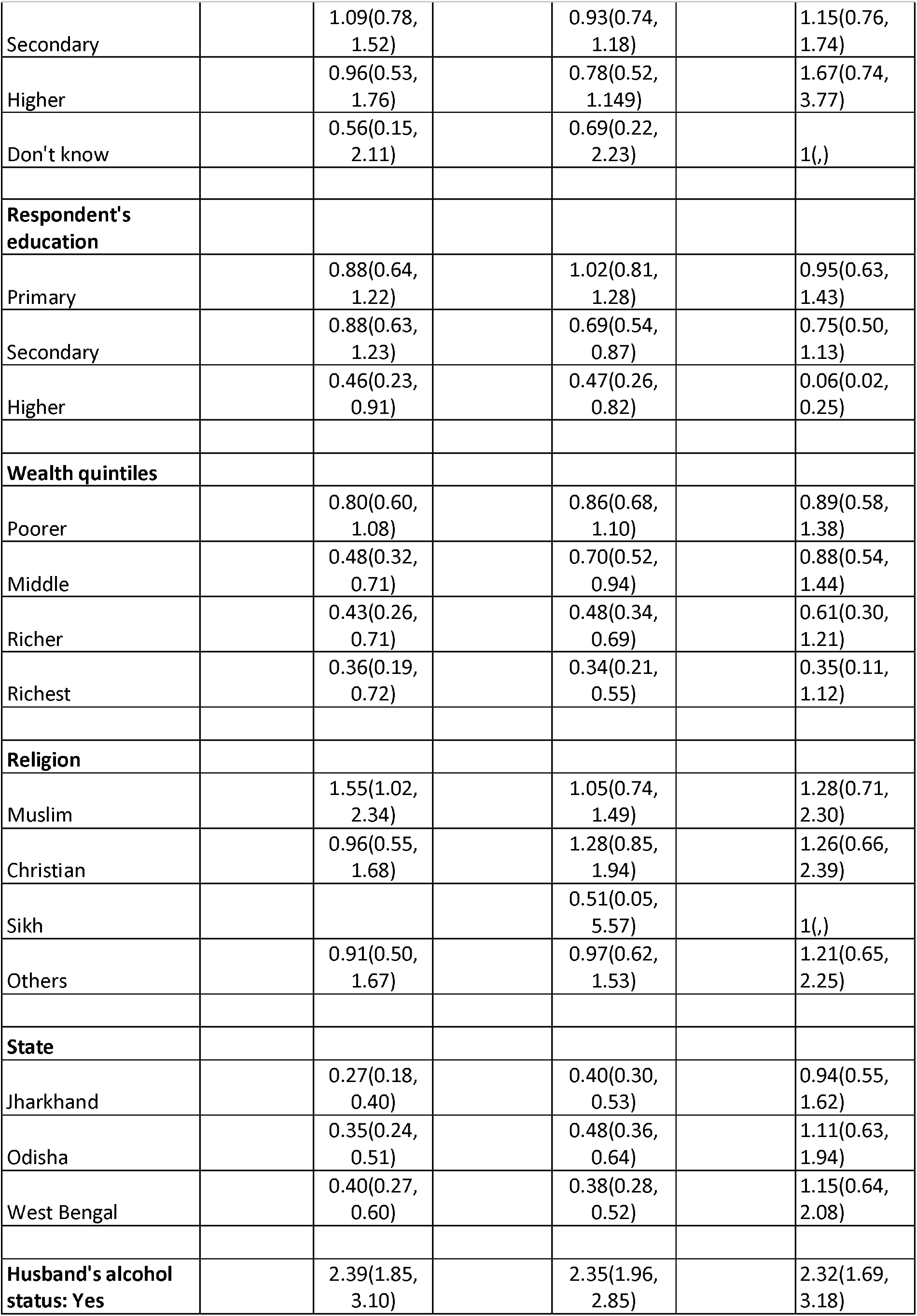
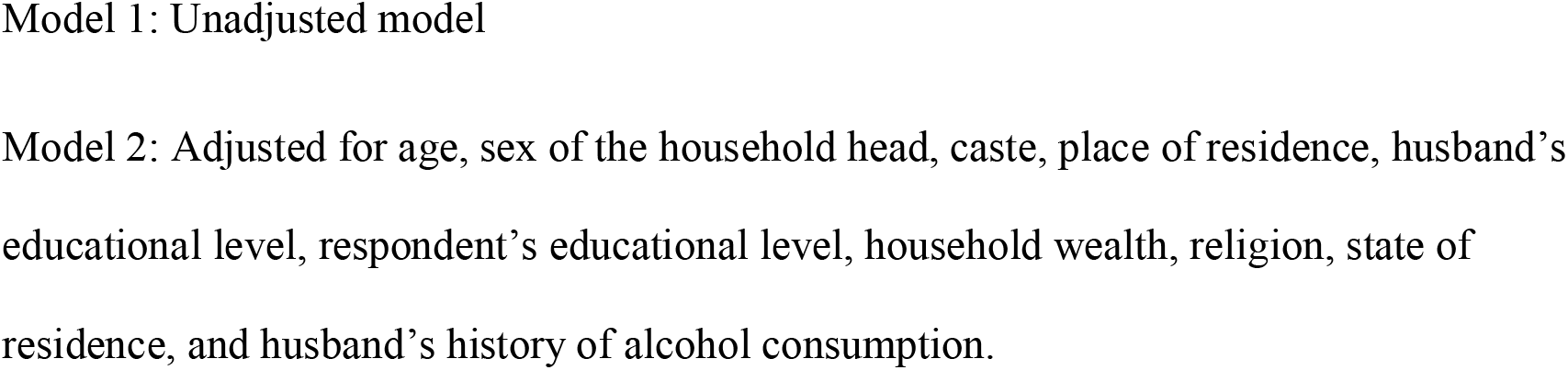
Association (odds ratios, 95% confidence interval) of exposure to cyclone with emotional, physical, and sexual IPV among ever married women in India.

Similarly, in fully adjusted models’ women with higher education attainment had lower odds of facing all forms of IPV compared to women who had no educational attainment. Further, in all adjusted models, husbands’ history of alcohol consumption had statistically significant positive association with all forms of IPV.

### 3.3. Exposure to drought and IPV

Neither single nor multivariable models found any significant association of exposure to drought with the three forms of IPV (see Table 4).

**Table 4:**
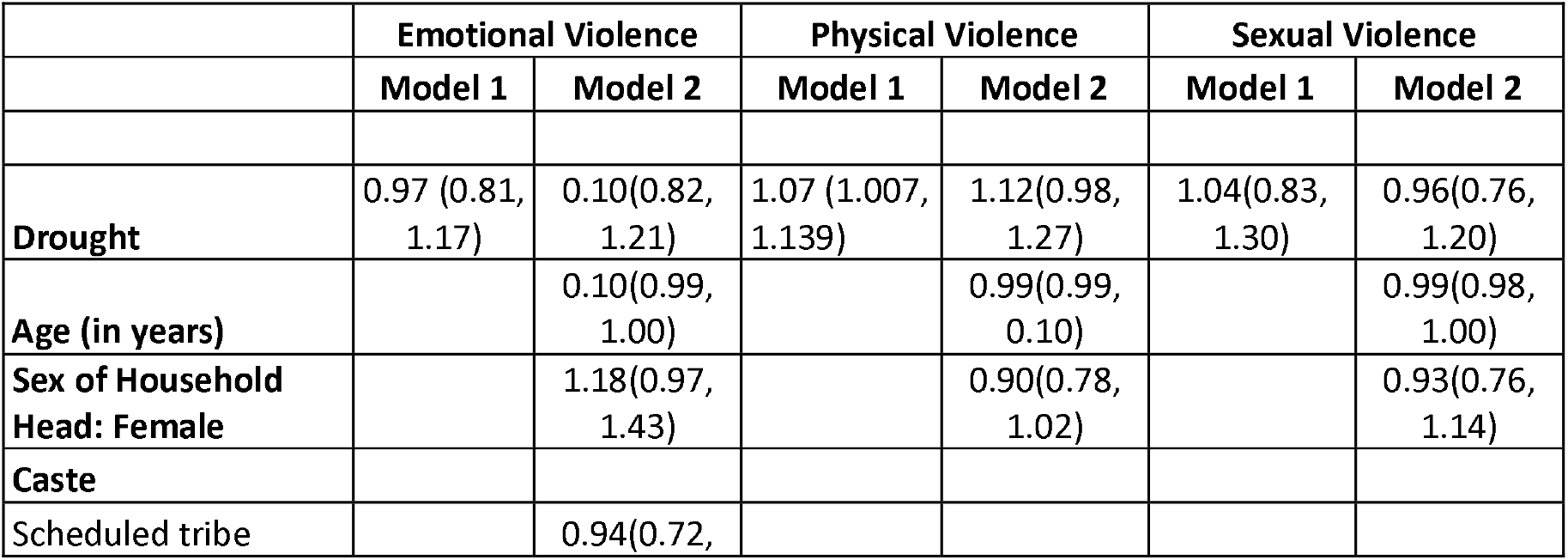
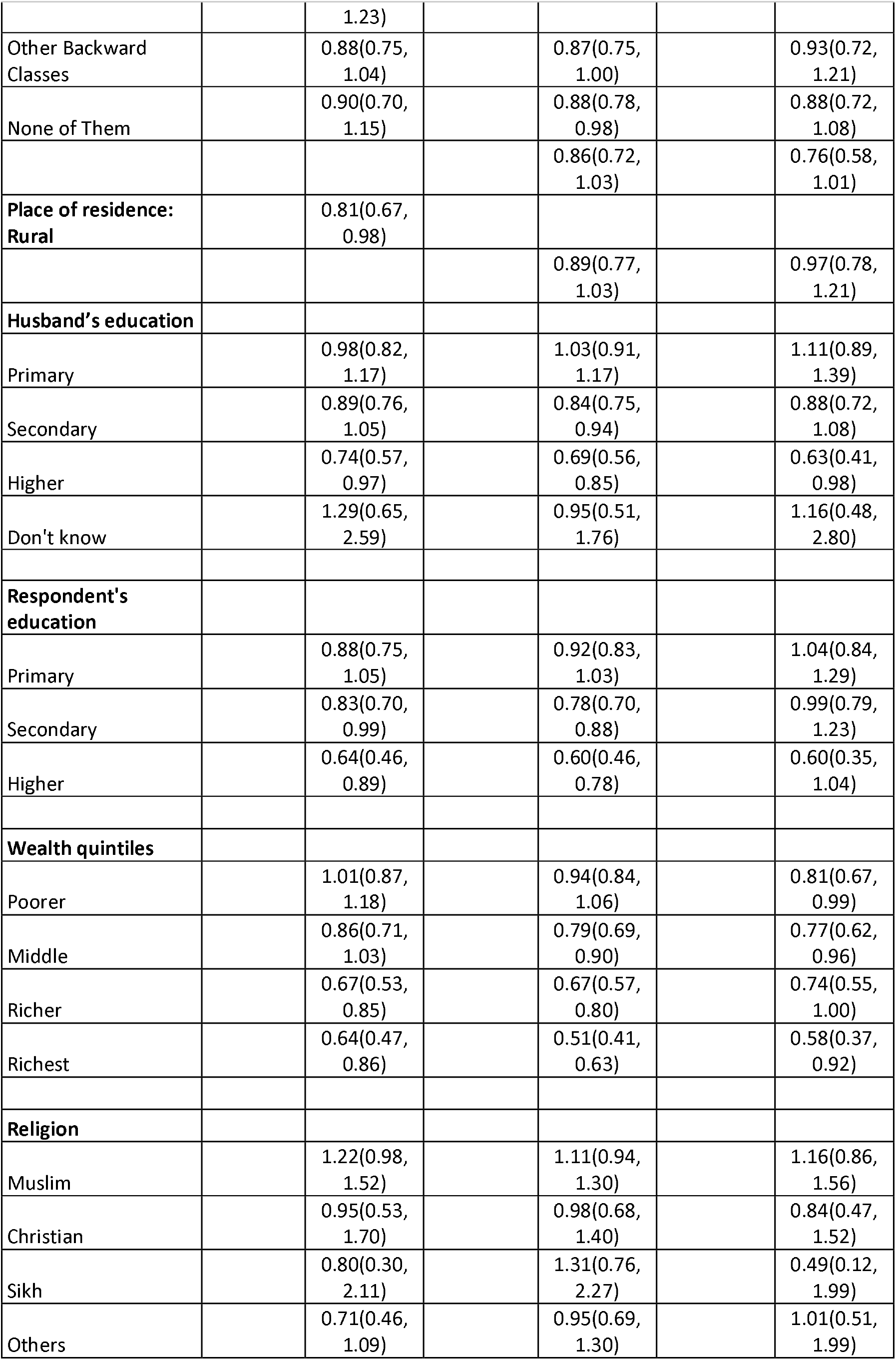
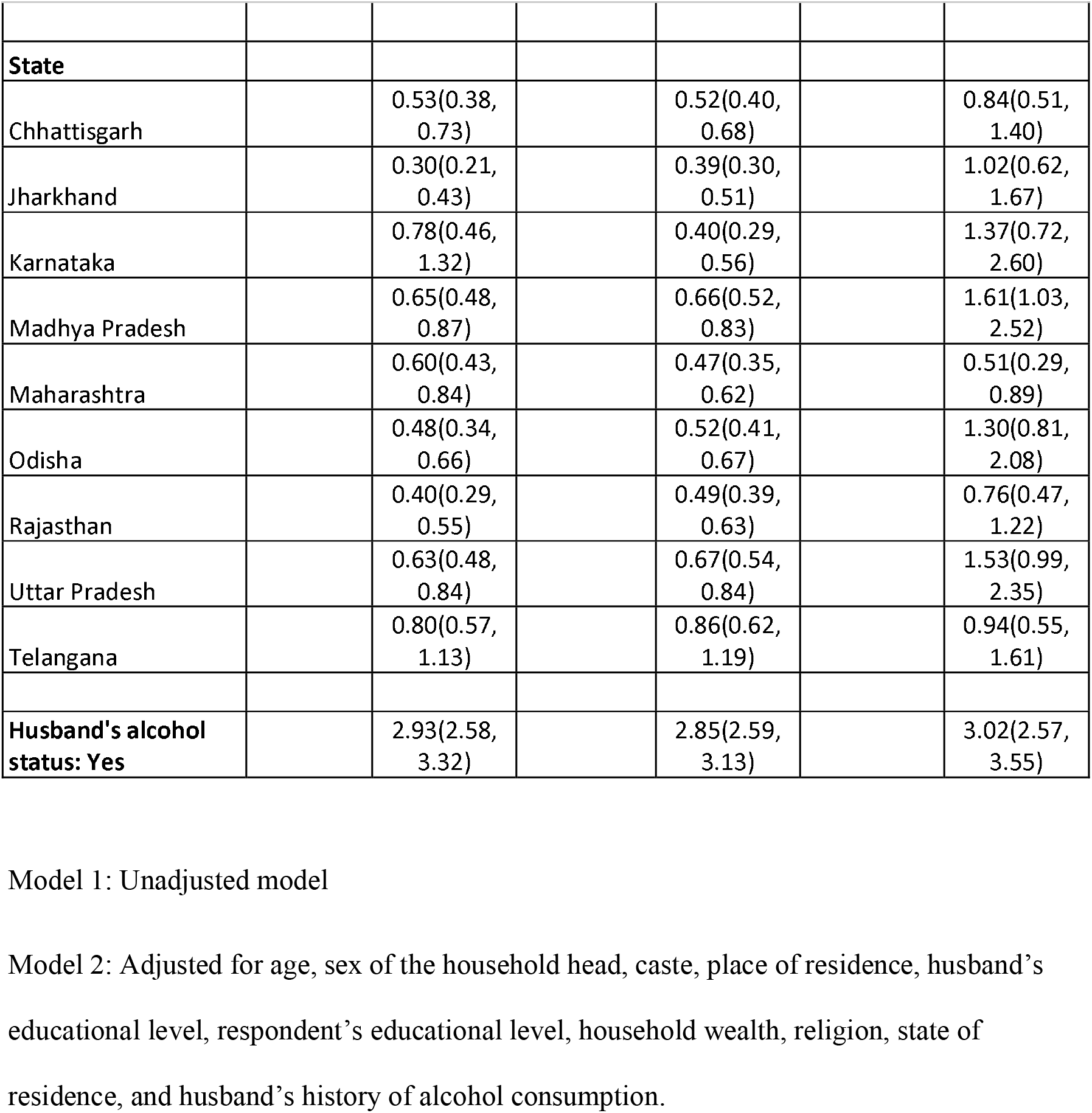
Association (odds ratios, 95% confidence interval) of exposure to drought with emotional, physical, and sexual IPV among ever married women in India.

Although not statistically significant, the direction of the association of exposure to drought with physical form of IPV was consistent with our hypothesis. Further, similar to the findings from the cyclone analyses, our fully adjusted models showed that women with higher educational attainment (compared to women with no educational attainment), or belonged to wealthier households (richer and richest compared to poorest households) had lower odds of reporting all forms of IPV. Additionally, all adjusted statistical models showed that partner’s history of alcohol consumption was significantly associated positively with all forms of IPV.

## 4. Discussion

### 4.1. Findings

Our analysis of data for four Indian states from the fourth National Family and Health Survey of India (NFHS-4) found a positive association of exposure to cyclone with three forms of IPV among ever-married Indian women. Consistent with our hypotheses, these findings suggest that Indian women may be at the risk of facing IPV in the aftermath of natural disasters. However, our findings did not support our hypothesis that exposure to drought would have a positive association with IPV. Further, women who belonged to wealthier households, women who were educated, and women whose husbands had no history of alcohol consumption, were less likely to experience any form of IPV, corroborating findings from previous Indian studies (33).

We found that women were at higher risk of emotional violence such as verbal abuse, humiliation, and threat to harm from intimate partners during the aftermath of a cyclone, a sudden-onset disaster. This corroborates findings from previous studies investigating violence against women after sudden-onset disasters such as floods (34), earthquakes (26), tsunamis (5,8,11,12,35), and cyclones (36). Cyclone Phailin hit the eastern coast of India on October 4, 2013, affecting the three Indian states of Odisha, Andhra Pradesh, and Jharkhand. According to EM-DAT (36), 13,230,000 people were affected, and the estimated cost of total damage was $633,471. On the other hand, cyclone Hudhud made its landfall on October 12, 2014, disrupting the lives of people in Andhra Pradesh, Odisha, and West Bengal. The estimated total damage from Hudhud was $7,000,000, affecting the lives of as many as 920,000 persons (37). Evidently, such sudden-onset disaster disrupted everyday lives and likely caused enormous financial crises within households, leading to adverse psychological outcomes among the household members. In patriarchal societies such as India, men are understood to have control over the wealth and household decision-making. Patriarchy also drives the norm that men act as providers who do not give into emotions. Thus, a failing household might convince men to believe that they lost their ‘provider and protector’ roles in supporting their families (especially the women and children), giving rise to emotional stress and depression among men. Such feelings of inadequacy may cause men to cope by using alcohol, gambling, and aggression, which might further lead them to abuse their wives, which is known to be used as a way to relieve stress in the Indian context (38,39). Previous research highlighted the effect of such post-traumatic stress on IPV perpetration (40,41). Moreover, different forms of hyper-masculinity, to compensate their loss of “protector/provider roles, could also emerge among men from their loss and ensuing stress, leading to increased levels of violence and discord in heterosexual relationships (1). This is likely to be observed in societies where women are perceived as subordinates to men. For instance, in countries like India, Pakistan, Cambodia, Mexico, Nigeria, and Tanzania, studies find that violence is often perceived as ‘physical chastisement’ i.e. a “husband’s right to correct an erring wife (1).” Generally speaking, such societies link masculinity with ‘toughness, male honour, or dominance’ and frequently accept the notion that men “own” women (42). Thus, women of their household might be easy targets for men to vent their anger on in such societies.

Research suggests that emotional abuse is significantly related to physical aggression (43). Emotional abuse was reported as a long-term predictor of physical violence among married couples in the United States (29). Thus, even though we found a statistically significant association of exposure to cyclone only with emotional violence, we argue that the women are more likely to face physical and sexual violence as well in the aftermath of cyclone. Notably, although not statistically significant, our results also support this conclusion. Our small sample size likely limited the statistical power and thus prevented us from finding statistically significant associations for physical and sexual violence.

On the other hand, our drought analysis revealed a mixed picture. Although not statistically significant, we found that exposure to drought had a positive relationship with physical IPV among Indian women. This is similar to the results of a previous study highlighting the relationship between drought and increased IPV in 19 sub-Saharan African countries (44). Drought affects agricultural productivity (45–47), and limits employment opportunities, which in turn could take a toll on the household income. Such financial instability in households could lead to frustration which may exacerbate mental processes leading to aggression. Insufficient income to meet the needs of one’s family also causes stress which could influence the degree of domestic violence. Further, financial strain in the household could disempower women, making them more dependent on their husbands, thus narrowing their opportunities to leave their abusive husbands. Previous studies have shown that income shocks could influence IPV in poorer households (29). Our study found statistically significant results to support these findings. We found that women belonging to poorer households are more likely to face domestic violence compared to those from wealthier households. Women lacking sufficient material resources to cope with natural disasters are likely to be more vulnerable to the effects of extreme weather events. Our findings, in the case of both drought and cyclone, corroborate this. The explanations for this could be that with greater financial resources, women might have higher levels of confidence and greater self-assurance, leading to an enhanced ability to manage relationships, prevent violent flare-ups, and an ability to separate from the spouse if needed (which may act as a deterrent to the perpetrator).

Exposure to drought has also been shown to increase sexually coercive behaviour towards women (29). However, we did not find evidence to support the associations of exposure to drought with emotional and sexual forms of IPV. There could be several reasons for this. First, the time of collection of NFHS-4 data coincided with the period of drought. Thus, several responses to questions measuring the outcomes might not have reflected the effect of the exposure, limiting our ability to accurately analyse the hypothesized relationship. For instance, the respondents who were surveyed in the initial months (January 2015 onwards) might not have given the same answer 4 to 5 months later, when the stress from drought may have built up. The impact of droughts likely manifests gradually, and as such, the respondents interviewed might have not experienced violence in the period when they were surveyed but may have later been inflicted by domestic violence. Second, emotional violence was more broadly defined than the other forms of violence in the DHS surveys (48) while questions on sexual violence risked social desirability bias, thus leading to a possible measurement error. These could jeopardise the accuracy of statistical relationships. However, even though our cyclone analysis followed the same definitions of IPV, we were able to show a statistically significant association of cyclone with emotional IPV. This suggests that the cross-sectional association of drought with IPV is a tenuous one.

### 4.2. Limitations and strengths

There are several limitations of this study that we acknowledge. First, the cross-sectional nature of the survey limits our ability to claim causal effects. However, given the very less probability of our outcome causing the exposure, we argue that our hypothesized direction of the relationships is supported. Second, our analyses were performed with district-level exposures, assuming that all households within the districts labelled as exposed had indeed been uniformly exposed to the cyclones/drought. Practically, this might not be the case, as a few households in the exposed districts might have not been affected at all by either of the disasters. However, to our knowledge this is the first study from India to operationalize exposure to natural disasters at the district level, which strengthens the assignment of the exposure as compared to previous studies which have assigned exposure at the state level.

Third, our sample included only women who were either married, thus, limiting the generalizability of our findings to women with other relationship statuses. However, given that we found important findings in a sample of women in a socially sanctioned union, our findings suggest that the women who are in socially-stigmatized relationships could be at higher risk of IPV, due to a greater pressure in concealing their relationship intact. We therefore argue that the risk of post-disaster IPV in such women in at least the same as that of married women. Moreover, we analysed a sample representing most women in romantic relationships in India. Thus, only a small portion of women are underrepresented in our study. Fourth, the measures of IPV relied on self-reported data from the women experiencing it, suggesting possible respondent bias (4). However, since around 52% of the women in the national sample justified IPV based on at least one reason (17), we argue that then social desirability pressure to hide IPV is not high.

Despite these limitations, the major strength of the study is that it drew from a sample that is representative of all the states which experienced either the drought or the cyclones under study to analyse the relationship between two different types of natural disasters and three different forms of intimate partner violence. Our study restricts the comparison of women residing in exposed districts to women residing in unexposed districts only in those states which experience the natural disasters under study, thus setting up a conservative test of the hypothesis unlike a state-level exposure assignment. Further, the study performed a district-level analysis, to understand the within-state variations in the exposure that had an impact on different forms of IPV. Thus, our findings provide evidence to formulate district-level disaster response measures. Our study found the association of natural disasters also highlights the long-term impact of natural disasters, the need for timely and appropriate data collection during the process of rebuilding post-disasters, to allow accurate investigations of the magnitude of this relationship.

## 5. Implications and conclusion

Our study contributes to the growing global literature on the public health impact of natural disasters. It highlights the potential of natural disasters in increasing the risk of IPV and thus worsen the psychological and physical well-being of IPV survivors when they are already suffering from the impact of the disaster. Our findings also echo what Shultz et al. (49) quoted: “The psychological footprint of disaster is larger than the medical footprint.” Thus, they highlight the need for disaster relief efforts to be context- and gender-sensitive. (50) IPV magnifies the burden borne by women during and after any disaster. However, globally, the post-disaster government policies have often failed to integrate the risks from these two sources. Disaster preparedness and response programs have considered the social context to a limited extent (35). For instance, in Haiti, after the 2010 earthquake women were forced to grant sexual favours to obtain basic needs and access to supplies (35), a fact missed by several response teams. Policymakers could use our findings to design socially inclusive disaster-recovery programs. For instance, the emergency response team could be gender sensitized and trained using role-playing (and other methods) to perceive subtle signs of IPV and share IPV-prevention messages during disaster times. Mobile applications designed to link disaster survivors with response teams could also have a built-in method of safely reporting IPV. These efforts could be seamlessly integrated with non-governmental organizations involved in disaster relief, thus further enhancing their current efforts to spread awareness about gender-equal attitudes in the local communities. Additionally, in order to address the psychological burden of surviving disasters in conjunction with being an IPV survivor disaster relief programs could include a counselling component. Qualitative studies unpacking the mechanisms linking natural disasters with IPV and longitudinal studies investigating the lag effects of disasters in detail could shed more light on this complex issue.

All in all, given the increased frequency of natural disasters expected due to climate change, India needs to prepare for the social disasters that they might bring.

## Data Availability

The study used secondary data (National Family Health Survey-4, India) for the analyses.

https://dhsprogram.com/what-we-do/survey/survey-display-355.cfm

## Acknowledgement

AJS has received a fellowship by Department of Science and Technology, India while working on this project.

